# Efficacy and Safety of SGLT2 Inhibitors in the Treatment of Type 2 Diabetes: An Umbrella Review

**DOI:** 10.1101/2025.11.27.25341177

**Authors:** Liu Haoling, Rui Ma

## Abstract

**Objective:** While sodium-glucose co-transporter 2 (SGLT2) inhibitors offer a novel, insulin-independent approach to managing type 2 diabetes (T2DM), their overall benefit-risk profile, encompassing cardio-renal outcomes and long-term safety, requires a comprehensive synthesis of the evidence. This umbrella review aims to definitively evaluate the efficacy and safety of SGLT2 inhibitors in patients with T2DM.

**Methods:** This umbrella review systematically searched major databases for relevant systematic reviews and meta-analyses up to September 2025. The methodological quality and certainty of evidence were assessed using AMSTAR 2 and GRADE tools.

**Results:** SGLT2 inhibitors demonstrated significant benefits in glycemic control (HbA1c WMD: −0.52% to −0.56%), body weight (MD: −1.76 to −2.63 kg), and systolic blood pressure (WMD: −4.08 mmHg). They also showed marked cardio-renal protection, reducing risks of major adverse cardiovascular events (RR=0.85), hospitalization for heart failure (RR=0.67), cardiovascular death (RR=0.75), all-cause mortality (RR=0.79), and composite renal outcomes (RR=0.59–0.64). Additionally, they positively modulated inflammatory markers and adipokines. However, these benefits were counterbalanced by increased risks of genital infections (OR=3.57), urinary tract infections (OR=1.34), and diabetic ketoacidosis (OR=2.19). The overall quality of evidence was generally low to very low.

**Conclusion:** SGLT2 inhibitors offer a multi-faceted therapeutic option for T2DM, providing glycemic, cardiovascular, and renal benefits, which make them particularly valuable for high-risk patients. Clinicians should be aware of the associated adverse events. Future high-quality, long-term studies are warranted to strengthen these findings.

---

The management of hyperglycemia in type 2 diabetes mellitus (T2DM) remains challenging due to the limited efficacy and adverse side effects of traditional therapies. As a result, the development of novel glucose-lowering drugs has attracted increasing attention, among which sodium-glucose co-transporter 2 inhibitors (SGLT2 inhibitors) have emerged as a highly promising class of agents^1–6^. By inhibiting glucose reabsorption in the proximal renal tubules, SGLT2 inhibitors promote urinary glucose excretion and improve glycemic control, independent of insulin secretion or sensitivity^7–11^. In addition to their glucose-lowering effects, SGLT2 inhibitors offer multiple benefits, including weight reduction and blood pressure lowering, which has led to their increasing use in combination with metformin as part of dual therapy^10,12–14^. Furthermore, growing evidence suggests that these agents provide cardiovascular and renal protection in high-risk T2DM patients. However, the precise extent of these benefits—particularly their impact on hard renal endpoints such as kidney failure, dialysis, transplantation, or death due to kidney disease—remains to be fully elucidated, as previous studies have not adequately assessed outcome differences across various stages of estimated glomerular filtration rate (eGFR) and proteinuria^14–17^. Recent observations also suggest that SGLT2 inhibitors may enhance insulin sensitivity and modulate inflammatory biomarkers, mechanisms that are closely related to the pathophysiological processes of diabetes and its cardio-renal complications^13,18–20^. In addition, although these drugs are generally well tolerated, the risk of adverse reactions still needs to be further explored in real-world studies^8,14,21,22^. The currently approved SGLT2 inhibitors differ in pharmacological potency—some inhibit only renal glucose transporters, while others act on both renal and intestinal transporters—thus, comparative studies are needed to assess their efficacy and safety^11,18,19,23^. Therefore, a comprehensive synthesis of existing evidence through systematic reviews and meta-analyses is essential to clarify the multiple effects of SGLT2 inhibitors on glycemic control, cardiovascular and renal outcomes, inflammatory markers, and safety, thereby providing guidance for clinical practice in type 2 diabetes^17,19,24,25^.

As a higher-level method of evidence synthesis, umbrella review integrates existing systematic reviews and meta-analyses to evaluate the strength of evidence for different associations using unified standards, identify potential biases, and comprehensively determine the effectiveness of SGLT2 inhibitors on various health outcomes^26–29^. Based on this, the present study adopts the umbrella review approach, strictly following the PRISMA-P guidelines, to systematically integrate published evidence from relevant systematic reviews and meta-analyses^30,31^. The aim is to comprehensively assess the effects of SGLT2 inhibitors on multiple health indicators, including blood glucose control, cardiovascular outcomes, renal function, weight changes, and safety, to clarify the evidence levels for different clinical populations and endpoint outcomes, provide a scientific basis for the clinical application and individualized treatment strategies of SGLT2 inhibitors, and offer a reference for future research directions^11,14,16,32^.

## 1 Methods

### 1. Umbrella Review Method

We systematically collected and integrated data from multiple systematic reviews (SRs) and meta-analyses (MAs) on the impact of Sodium-Glucose Transporter 2 Inhibitors interventions on health outcomes. By comprehensively evaluating all relevant clinical outcome information, we aimed to outline the full scope of evidence in this field and provide integrated insights for the application of SGLT2 interventions in healthcare. The protocol for this umbrella review was pre-registered on PROSPERO (ID: CRD420251145145), and the study results are reported in accordance with the Preferred Reporting Items for Systematic Reviews and Meta-Analyses (PRISMA) guidelines.

### 1.2 Literature Search

To comprehensively obtain relevant literature, we searched multiple authoritative databases, covering the period from their inception to August 2025, including but not limited to Embase, Medline, the Cochrane Database of Systematic Reviews, and Web of Science. A pre-designed search strategy was used: (Sodium-Glucose Transporter 2 Inhibitors) AND (systematic review or meta-analysis), strictly following the SIGN guidelines to ensure accuracy and comprehensiveness. In addition, we carefully reviewed the reference lists of all peer-reviewed articles that met the preliminary screening criteria to avoid missing any potentially relevant studies. Discrepancies in the literature screening process were resolved by a third professional researcher to reach a final consensus.

### 1.3 Inclusion and Exclusion Criteria

This study included systematic reviews and meta-analyses of observational and/or interventional studies evaluating the application of Sodium-Glucose Transporter 2 Inhibitors as an intervention in the field of human health. Participants included populations of any country or region, race, or gender. If a single article presented multiple health outcomes, each outcome was extracted and analyzed separately. When two or more meta-analyses addressed the same Sodium-Glucose Transporter 2 Inhibitors intervention topic, priority was given to the meta-analysis with a broader study population. In addition, data from meta-analyses with smaller sample sizes but without overlap with other studies were also extracted to maximize the use of all relevant information. We excluded literature not published in English, as well as literature based on animal experiments and/or in vitro study data. Furthermore, articles focusing solely on the principles of Sodium-Glucose Transporter 2 Inhibitors without assessing their impact on health outcomes were also excluded. If two or more meta-analyses existed on the same topic, priority was given to the meta-analysis with a broader study population.

### 1.4 Data Extraction

Data were extracted from the included articles by two independent researchers. The extracted data included: 1) name of the first author and year of publication; 2) journal name; 3) characteristics of the study population; 4) health outcome indicators; 5) interventions; 6) number of cases in each meta-analysis; 7) number of original studies included in each meta-analysis; 8) study design type of the original studies; 9) effect size measures used in the meta-analysis and their corresponding estimates; 13) type of effect model (note: numbering follows the original text); 10) results of heterogeneity tests; 11) results of publication bias assessment. For quantitative synthesis, priority was given to fully adjusted, study-specific pooled effect size estimates (including odds ratio [OR], mean estimate [ME], relative risk [RR], weighted mean difference [WMD], standardized mean difference [SMD], etc.) and their 95% confidence intervals (CIs). In case of discrepancies in data extraction, a third researcher reviewed the data and made the final decision.

### 1.5 Methodological Quality Assessment and Evidence Grading

The methodological quality of the included studies was rigorously assessed using the 11 items of AMSTAR2 to ensure the scientific rigor and standardization of the research process^33^. At the same time, the GRADE system was used to grade the quality of evidence and provide corresponding recommendations, thereby clarifying the reliability and credibility of different research results^34^.

### 1.6 Data Analysis

For each meta-analysis, a random-effects model (or a fixed-effects model as actually applied) was used to present the extracted intervention exposure data, health outcome data, and fully adjusted study-specific pooled effect sizes with their 95% CIs. The I ^2^ statistic and Cochran’s Q test were used to assess the degree of heterogeneity between studies and to determine the consistency of the results. Egger’s test was used to assess small-study effects (marked as NA if not reported in the literature). For small-study effects and heterogeneity tests, a P-value < 0.1 was considered statistically significant; for other tests, a P-value < 0.05 was considered statistically significant.

## 2 Outcome

### 2.1 Characteristics of MA

Figure 1 shows the flowchart of the literature selection process. After a systematic search, a total of 5759 articles were identified. After applying the inclusion and exclusion criteria, 36 MAs were included, covering 8 different outcome measures (Figure 2). The characteristics of all the included studies are shown in Table 1.

**Fig. 1.**
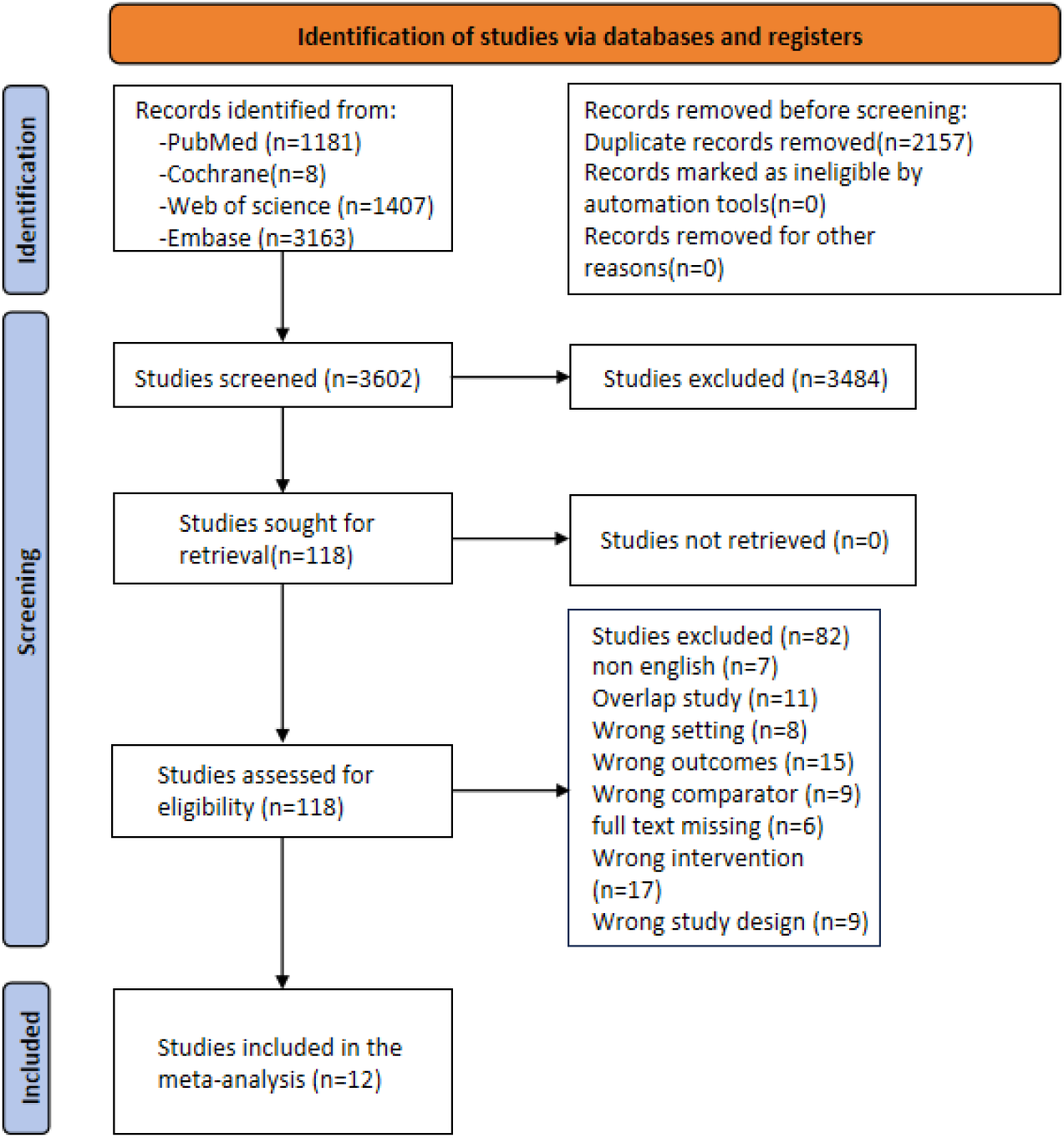
Flowchart of the selection process.

**Figure 2.**
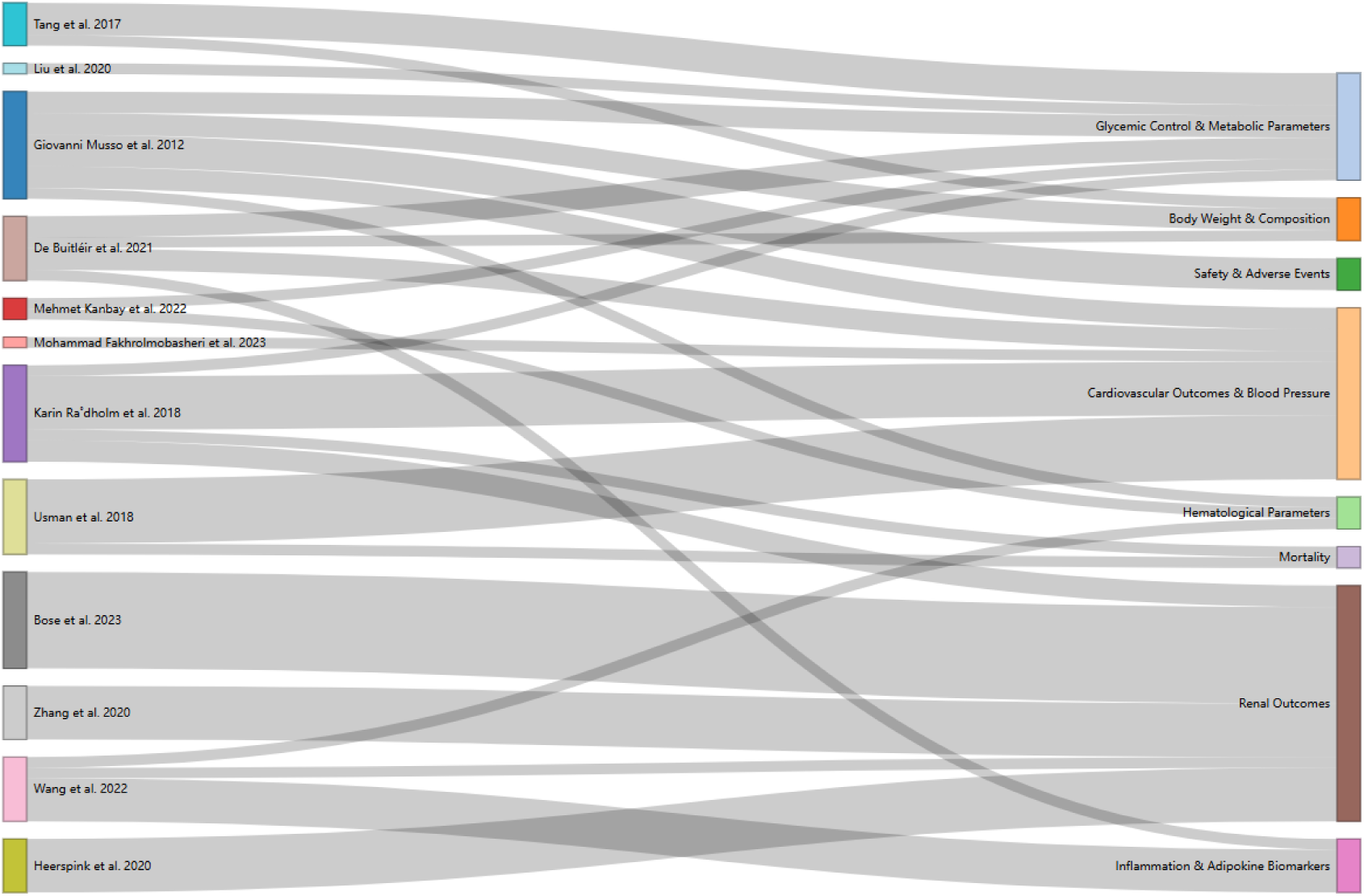
Sankey diagram showing the associations between various studies and health-related outcome categories

**Table 1.** Evidence summary.

| Outcome | Intervention measures for the experimental group | Intervention measures for the control group | Study | No. of total | group | No. of studies in MA | RCT | Cohort | Case control | Effects model | I<sup>2</sup> | Egger test P value |
| --- | --- | --- | --- | --- | --- | --- | --- | --- | --- | --- | --- | --- |
| Glycated Hemoglobin | Sodium glucose co-transport-2 inhibitors | Placebo | Giovanni Musso et al. 2012<sup>7</sup> | 4063 | Type 2 Diabetes Mellitus | 13 | 13 | 0 | 0 | Fixed | 0.14 | NA |
| Fasting Plasma Glucose | Sodium glucose co-transport-2 inhibitors | Placebo | Giovanni Musso et al. 2012<sup>7</sup> | 4063 | Type 2 Diabetes Mellitus | 13 | 13 | 0 | 0 | Fixed | 0.18 | NA |
| Body Mass Index | Sodium glucose co-transport-2 inhibitors | Placebo | Giovanni Musso et al. 2012<sup>7</sup> | 4063 | Type 2 Diabetes Mellitus | 13 | 13 | 0 | 0 | Random | 0.9 | NA |
| Waist Circumference | Sodium glucose co-transport-2 inhibitors | Placebo | Giovanni Musso et al. 2012<sup>7</sup> | 4063 | Type 2 Diabetes Mellitus | 13 | 13 | 0 | 0 | Fixed | 0 | NA |
| Systolic Blood Pressure | Sodium glucose co-transport-2 inhibitors | Placebo | Giovanni Musso et al. 2012<sup>7</sup> | 4063 | Type 2 Diabetes Mellitus | 13 | 13 | 0 | 0 | Fixed | 0 | NA |
| Diastolic Blood Pressure | Sodium glucose co-transport-2 inhibitors | Placebo | Giovanni Musso et al. 2012<sup>7</sup> | 4063 | Type 2 Diabetes Mellitus | 13 | 13 | 0 | 0 | Fixed | 0 | NA |
| Serum Uric Acid | Sodium glucose co-transport-2 inhibitors | Placebo | Giovanni Musso et al. 2012<sup>7</sup> | 4063 | Type 2 Diabetes Mellitus | 13 | 13 | 0 | 0 | Random | 0.5 | NA |
| Incident Hypoglycemia | Sodium glucose co-transport-2 inhibitors | Placebo | Giovanni Musso et al. 2012<sup>7</sup> | 4063 | Type 2 Diabetes Mellitus | 13 | 13 | 0 | 0 | Fixed | 0 | NA |
| Urinary Tract Infection | Sodium glucose co-transport-2 inhibitors | Placebo | Giovanni Musso et al. 2012<sup>7</sup> | 4063 | Type 2 Diabetes Mellitus | 13 | 13 | 0 | 0 | Fixed | 0 | NA |
| Genital Tract Infection | Sodium glucose co-transport-2 inhibitors | Placebo | Giovanni Musso et al. 2012<sup>7</sup> | 4063 | Type 2 Diabetes Mellitus | 13 | 13 | 0 | 0 | Fixed | 0 | NA |
| Hemoglobin Level | Sodium-glucose cotransporter 2 inhibitors | Placebo | Mehmet Kanbay et al. 2022<sup>9</sup> | 14748 | Type 2 Diabetes Mellitus | 7 | 7 | 0 | 0 | Random | 0.94 | NA |
| Hematocrit Level | Sodium-glucose cotransporter 2 inhibitors | Placebo | Mehmet Kanbay et al. 2022<sup>9</sup> | 14748 | Type 2 Diabetes Mellitus | 13 | 13 | 0 | 0 | Random | 0.99 | NA |
| Insulin Sensitivity/Insulin Resistance in T2DM Population | Sodium-glucose cotransporter 2 inhibitors | Placebo | Mohammad Fakhrolmobasheri et al. 2023<sup>19</sup> | 1178 | Type 2 Diabetes Mellitus | 13 | 13 | 0 | 0 | Random | 0.848 | 0.19 |
| Major Cardiovascular Events | Sodium-glucose cotransporter 2 inhibitors | Placebo/active control | Karin Ra°dholm et al. 2018<sup>15</sup> | 32893 | Type 2 Diabetes Mellitus | 92 | 82 | 0 | 0 | Fixed | 0 | NA |
| Cardiovascular Death | Sodium-glucose cotransporter 2 inhibitors | Placebo | Karin Ra°dholm et al. 2018<sup>15</sup> | 30210 | Type 2 Diabetes Mellitus | 92 | 82 | 0 | 0 | Fixed | 0.804 | NA |
| Non-fatal Myocardial Infarction | Sodium-glucose cotransporter<br>2 inhibitors | Placebo | Karin Ra°dholm et al.<br>2018<sup>15</sup> | 30210 | Type 2<br>Diabetes<br>Mellitus | 92 | 82 | 0 | 0 | Fixed | 0 | NA |
| Non-fatal Stroke | Sodium-glucose cotransporter<br>2 inhibitors | Placebo | Karin Ra°dholm et al.<br>2018<sup>15</sup> | 30210 | Type 2<br>Diabetes<br>Mellitus | 92 | 82 | 0 | 0 | Fixed | 0.614 | NA |
| Hospitalisation for Unstable Angina | Sodium-glucose cotransporter<br>2 inhibitors | Placebo | Karin Ra°dholm et al.<br>2018<sup>15</sup> | 30210 | Type 2<br>Diabetes<br>Mellitus | 92 | 82 | 0 | 0 | Fixed | 0 | NA |
| Hospitalisation for Heart Failure | Sodium-glucose cotransporter<br>2 inhibitors | Placebo | Karin Ra°dholm et al.<br>2018<sup>15</sup> | 30210 | Type 2<br>Diabetes<br>Mellitus | 92 | 82 | 0 | 0 | Fixed | 0 | NA |
| All-cause Death | Sodium-glucose cotransporter<br>2 inhibitors | Placebo | Karin Ra°dholm et al.<br>2018<sup>15</sup> | 30210 | Type 2<br>Diabetes<br>Mellitus | 92 | 82 | 0 | 0 | Fixed | 0.546 | NA |
| Composite renal outcomes | Sodium-glucose cotransporter<br>2 inhibitors | Placebo | Karin Ra°dholm et al.<br>2018<sup>15</sup> | 18064 | Type 2<br>Diabetes<br>Mellitus | 92 | 82 | 0 | 0 | Fixed | 0 | NA |
| Albuminuria Progression | Sodium-glucose cotransporter<br>2 inhibitors | Placebo | Karin Ra°dholm et al.<br>2018<sup>15</sup> | 15139 | Type 2<br>Diabetes<br>Mellitus | 92 | 82 | 0 | 0 | Fixed | 0 | NA |
| Change in HbA<sub>1c</sub> from baseline | Sodium-glucose<br>cotransporter-2 inhibitors | Placebo | De Buitléir et al. 2021<sup>35</sup> | 1661 | Type 2<br>Diabetes<br>Mellitus | 6 | 6 | 0 | 0 | Random | 0.79 | NA |
| Change in fasting plasma glucose from<br>baseline | Sodium-glucose<br>cotransporter-2 inhibitors | Placebo | De Buitléir et al. 2021<sup>35</sup> | 1661 | Type 2<br>Diabetes<br>Mellitus | 6 | 6 | 0 | 0 | Random | 0 | NA |
| Change in weight from baseline | Sodium-glucose cotransporter-2 inhibitors | Placebo | De Buitléir et al. 2021<sup>35</sup> | 1661 | Type 2 Diabetes Mellitus | 6 | 6 | 0 | 0 | Random | 0 | NA |
| Change in systolic blood pressure from baseline | Sodium-glucose cotransporter-2 inhibitors | Placebo | De Buitléir et al. 2021<sup>35</sup> | 1661 | Type 2 Diabetes Mellitus | 6 | 6 | 0 | 0 | Random | 0 | NA |
| Change in diastolic blood pressure from baseline | Sodium-glucose cotransporter-2 inhibitors | Placebo | De Buitléir et al. 2021<sup>35</sup> | 1661 | Type 2 Diabetes Mellitus | 4 | 4 | 0 | 0 | Random | 0 | NA |
| Proportion of participants achieving HbA<sub>1c</sub> | Sodium-glucose cotransporter-2 inhibitors | Placebo | De Buitléir et al. 2021<sup>35</sup> | 1661 | Type 2 Diabetes Mellitus | 6 | 6 | 0 | 0 | Random | 0.2 | NA |
| Change in C-reactive protein (CRP) from baseline | Sodium-glucose cotransporter-2 inhibitors | Placebo | Wang et al. 2022<sup>36</sup> | 6261 | Type 2 Diabetes Mellits | 14 | 14 | 0 | 0 | Random | 0.5 | NA |
| Change in tumor necrosis factor-alpha (TNF-α) from baseline | Sodium-glucose cotransporter-2 inhibitors | Placebo | Wang et al. 2022<sup>36</sup> | 6261 | Type 2 Diabetes Mellitus | 7 | 7 | 0 | 0 | Random | 0 | NA |
| Change in interleukin-6 (IL-6) from baseline | Sodium-glucose cotransporter-2 inhibitors | Placebo | Wang et al. 2022<sup>36</sup> | 6261 | Type 2 Diabetes Mellitus | 6 | 6 | 0 | 0 | Random | 0.88 | NA |
| Change in leptin from baseline | Sodium-glucose cotransporter-2 inhibitors | Placebo | Wang et al. 2022<sup>36</sup> | 6261 | Type 2 Diabetes Mellitus | 9 | 9 | 0 | 0 | Random | 0.6 | NA |
| Change in adiponectin from baseline | Sodium-glucose cotransporter-2 inhibitors | Placebo | Wang et al. 2022<sup>36</sup> | 6261 | Type 2 Diabetes Mellitus | 17 | 17 | 0 | 0 | Random | 0.26 | NA |
| Change in ferritin from baseline | Sodium-glucose cotransporter-2 inhibitors | Placebo | Wang et al. 2022<sup>36</sup> | 6261 | Type 2 Diabetes Mellitus | 6 | 6 | 0 | 0 | Random | 0.97 | NA |
| Composite renal outcome | Sodium-glucose cotransporter-2 inhibitors | Placebo | Bose et al. 2023<sup>37</sup> | 71553 | Type 2 Diabetes Mellitus | 9 | 9 | 0 | 0 | Random | 0.29 | NA |
| Decline in estimated glomerular filtration rate $\geq 40\%$ | Sodium-glucose cotransporter-2 inhibitors | Placebo | Bose et al. 2023<sup>37</sup> | 71553 | Type 2 Diabetes Mellitus | 4 | 4 | 0 | 0 | Random | 0.59 | NA |
| Doubling of serum creatinine | Sodium-glucose cotransporter-2 inhibitors | Placebo | Bose et al. 2023<sup>37</sup> | 71553 | Type 2 Diabetes Mellitus | 3 | 3 | 0 | 0 | Random | 0.34 | NA |
| Dialysis or renal replacement therapy | Sodium-glucose cotransporter-2 inhibitors | Placebo | Bose et al. 2023<sup>37</sup> | 71553 | Type 2 Diabetes Mellitus | 5 | 5 | 0 | 0 | Random | 0 | NA |
| Sustained estimated glomerular filtration rate $< 15$ ml/min/1.73m<sup>2</sup> for $\geq 30$ days | Sodium-glucose cotransporter-2 inhibitors | Placebo | Bose et al. 2023<sup>37</sup> | 71553 | Type 2 Diabetes Mellitus | 3 | 3 | 0 | 0 | Random | 0 | NA |
| End-stage renal disease | Sodium-glucose cotransporter-2 inhibitors | Placebo | Bose et al. 2023<sup>37</sup> | 71553 | Type 2 Diabetes Mellitus | 3 | 3 | 0 | 0 | Random | 0 | NA |
| Acute kidney injury | Sodium-glucose cotransporter-2 inhibitors | Placebo | Bose et al. 2023<sup>37</sup> | 71553 | Type 2 Diabetes Mellitus | 6 | 6 | 0 | 0 | Random | 0 | NA |
| Renal death | Sodium-glucose cotransporter-2 inhibitors | Placebo | Bose et al. 2023<sup>37</sup> | 71553 | Type 2 Diabetes Mellitus | 6 | 6 | 0 | 0 | Random | 0 | NA |
| Progression to macroalbuminuria | Sodium-glucose cotransporter-2 inhibitors | Placebo | Bose et al. 2023<sup>37</sup> | 71553 | Type 2 Diabetes Mellitus | 2 | 2 | 0 | 0 | Random | 0.91 | NA |
| Composite renal outcome | Sodium-glucose cotransporter-2 inhibitors | Placebo | Zhang et al. 2020<sup>38</sup> | 28529 | Type 2 Diabetes Mellitus | 3 | 3 | 0 | 0 | Random | 0.51 | NA |
| Acute renal failure or injury | Sodium-glucose cotransporter-2 inhibitors | Placebo | Zhang et al. 2020<sup>38</sup> | 48731 | Type 2 Diabetes Mellitus | 15 | 15 | 0 | 0 | Random | 0 | 0.029 |
| Renal impairment | Sodium-glucose cotransporter-2 inhibitors | Placebo | Zhang et al. 2020<sup>38</sup> | 28865 | Type 2 Diabetes Mellitus | 17 | 17 | 0 | 0 | Random | 0 | NA |
| Change in estimated glomerular filtration rate | Sodium-glucose cotransporter-2 inhibitors | Placebo | Zhang et al. 2020<sup>38</sup> | 15556 | Type 2 Diabetes Mellitus | 46 | 46 | 0 | 0 | Random | 0.99 | NA |
| Change in urine albumin-creatinine ratio | Sodium-glucose cotransporter-2 inhibitors | Placebo | Zhang et al. 2020<sup>38</sup> | 10129 | Type 2 Diabetes Mellitus | 18 | 18 | 0 | 0 | Random | 0.98 | NA |
| Composite renal outcome | Sodium-glucose cotransporter-2 inhibitors | Placebo | Heerspink et al. 2020<sup>39</sup> | 38612 | Type 2 Diabetes Mellitus | 4 | 4 | 0 | 0 | Random | 0 | NA |
| Acute kidney injury | Sodium-glucose cotransporter-2 inhibitors | Placebo | Heerspink et al. 2020<sup>39</sup> | 38625 | Type 2 Diabetes Mellitus | 4 | 4 | 0 | 0 | Random | 0 | 0.42 |
| Progression to macroalbuminuria | Sodium-glucose cotransporter-2 inhibitors | Placebo | Heerspink et al. 2020<sup>39</sup> | 38598 | Type 2 Diabetes Mellitus | 4 | 4 | 0 | 0 | Random | 0 | NA |
| Decline in eGFR ≥ 40% | Sodium-glucose cotransporter-2 inhibitors | Placebo | Heerspink et al. 2020<sup>39</sup> | 38581 | Type 2 Diabetes Mellitus | 4 | 4 | 0 | 0 | Random | 0 | NA |
| End-stage kidney disease | Sodium-glucose cotransporter-2 inhibitors | Placebo | Heerspink et al. 2020<sup>39</sup> | 38671 | Type 2 Diabetes Mellitus | 4 | 4 | 0 | 0 | Random | 0 | NA |
| All-cause mortality | Sodium-glucose cotransporter-2 inhibitors | Placebo | Usman et al. 2018<sup>22</sup> | 34987 | Type 2 Diabetes Mellitus | 18 | 18 | 0 | 0 | Random | 0 | NA |
| Major adverse cardiac events | Sodium-glucose cotransporter-2 inhibitors | Placebo | Usman et al. 2018<sup>22</sup> | 34987 | Type 2 Diabetes Mellitus | 31 | 31 | 0 | 0 | Random | 0 | NA |
| Non-fatal myocardial infarction | Sodium-glucose cotransporter-2 inhibitors | Placebo | Usman et al. 2018<sup>22</sup> | 34987 | Type 2 Diabetes Mellitus | 27 | 27 | 0 | 0 | Random | 0 | NA |
| Heart failure/hospitalisation for heart failure | Sodium-glucose cotransporter-2 inhibitors | Placebo | Usman et al. 2018<sup>22</sup> | 34987 | Type 2 Diabetes Mellitus | 14 | 14 | 0 | 0 | Random | 0 | NA |
| Non-fatal stroke | Sodium-glucose cotransporter-2 inhibitors | Placebo | Usman et al. 2018<sup>22</sup> | 34987 | Type 2 Diabetes Mellitus | 28 | 28 | 0 | 0 | Random | 0 | NA |
| Atrial fibrillation | Sodium-glucose cotransporter-2 inhibitors | Placebo | Usman et al. 2018<sup>22</sup> | 34987 | Type 2 Diabetes Mellitus | 16 | 16 | 0 | 0 | Random | 0 | NA |
| Unstable angina | Sodium-glucose cotransporter-2 inhibitors | Placebo | Usman et al. 2018<sup>22</sup> | 34987 | Type 2 Diabetes Mellitus | 18 | 18 | 0 | 0 | Random | 0 | NA |
| Change in glycated hemoglobin (HbA1c, %) | Sodium-glucose cotransporter-2 inhibitors | Placebo | Tang et al. 2017<sup>40</sup> | 4235 | Type 2 Diabetes Mellitus | 7 | 7 | 0 | 0 | Random | 0.79 | >0.05 |
| Change in fasting plasma glucose (FPG, mmol/L) | Sodium-glucose cotransporter-2 inhibitors | Placebo | Tang et al. 2017<sup>40</sup> | 4235 | Type 2 Diabetes Mellitus | 7 | 7 | 0 | 0 | Random | 0.68 | >0.05 |
| Change in body weight (kg) | Sodium-glucose cotransporter-2 inhibitors | Placebo | Tang et al. 2017<sup>40</sup> | 4235 | Type 2 Diabetes Mellitus | 7 | 7 | 0 | 0 | Random | 0.75 | >0.05 |
| Change in insulin dose (IU) | Sodium-glucose cotransporter-2 inhibitors | Placebo | Tang et al. 2017<sup>40</sup> | 4235 | Type 2 Diabetes Mellitus | 6 | 6 | 0 | 0 | Random | 0.91 | <0.01 |
| Diabetic ketoacidosis | Sodium-glucose<br>co-transporter-2 inhibitors | Placebo | Liu et al. 2020<sup>24</sup> | 51701 | Type 2<br>Diabetes<br>Mellitus | 30 | 30 | 0 | 0 | Fixed | 10% | NA |

### 2.2 Blood Glucose Control and Metabolic Parameters

SGLT2 inhibitors demonstrated significant improvements in blood glucose control and metabolic parameters. In patients with type 2 diabetes, the weighted mean difference (WMD) in glycated hemoglobin (HbA₁c) levels was −0.56 (95% CI: −0.67 to −0.44); the WMD in fasting plasma glucose (FPG) was −18.28 mg/dL (95% CI: −20.66 to −15.89) (additionally, the WMD in change in FPG was −0.95 mmol/L, 95% CI: −1.21 to −0.70). The standardized mean difference (SMD) for insulin sensitivity/resistance was 0.72 (95% CI: 0.32 to 1.12). The odds ratio (OR) for achieving the HbA₁c target was 3.07 (95% CI: 2.29 to 4.12). The WMD for changes in insulin dosage was −8.79 IU (95% CI: −13.36 to −4.22). The odds ratio (OR) for the occurrence of diabetic ketoacidosis was 2.19 (95% CI: 1.41 to 3.39) (Figure 3).

**Figure 3:**
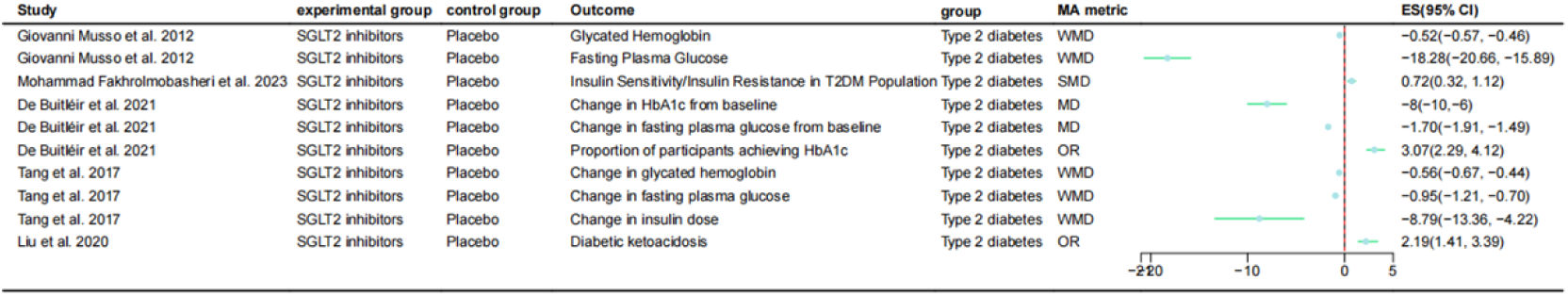
Forest Plot of the Effects of SGLT2 on Blood Glucose Control and Metabolic Parameters in Multiple Populations

### 2.3 Weight and Body Composition

SGLT2 inhibitors have a significant effect on improving body weight and body composition. In patients with type 2 diabetes, the weighted mean difference (WMD) in body mass index (BMI) was −1.17 (95% CI: −1.41 to −0.92); the weighted mean difference (WMD) in waist circumference was −1.20 cm (95% CI: −2.00 to −0.43). The mean difference (MD) in body weight change was −1.76 kg (95% CI: −2.04 to −1.48); the weighted mean difference (WMD) in body weight change was −2.63 (95% CI: −3.10 to −2.16) (Figure 4).

**Figure 4:**
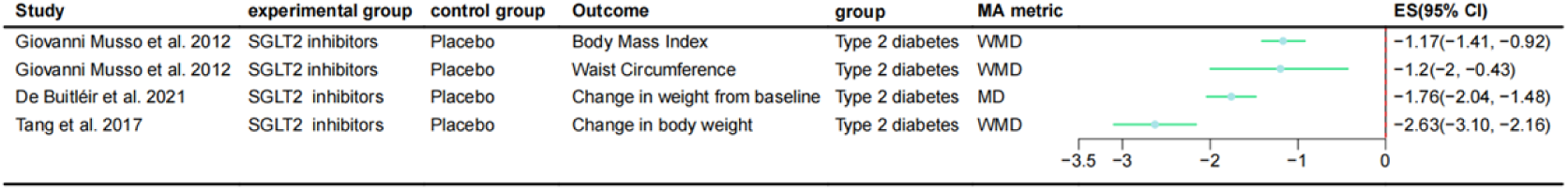
Forest Plot of the Effects of SGLT2 on Weight and Body Composition

### 2.4 Cardiovascular Outcomes and Blood Pressure

SGLT2 inhibitors have significant benefits in cardiovascular outcomes and blood pressure control. The weighted mean difference (WMD) in systolic blood pressure was −4.08 mmHg (95% CI: −4.91 to −3.24); the WMD in diastolic blood pressure was −1.16 mmHg (95% CI: −1.67 to −0.66) (additionally, the MD in change in SBP was −3.6 mmHg, 95% CI: −4.8 to −2.4; the MD in change in DBP was −1.5 mmHg, 95% CI: −2.4 to −0.6). The risk ratio (RR) for major cardiovascular events was 0.85 (95% CI: 0.77 to 0.93); the RR for cardiovascular death was 0.75 (95% CI: 0.65 to 0.87). The RR for nonfatal myocardial infarction was 0.84 (95% CI: 0.73 to 0.98); the RR for hospitalization for heart failure was 0.67 (95% CI: 0.55 to 0.80) (additionally, the OR for heart failure hospitalization was 0.67, 95% CI: 0.59 to 0.76). No significant improvement was observed in nonfatal stroke (RR = 1.03, 95% CI: 0.86 to 1.24; OR = 1.02, 95% CI: 0.85 to 1.21) and hospitalization for unstable angina (RR = 0.95, 95% CI: 0.73 to 1.24; OR = 0.95, 95% CI: 0.73 to 1.25) (Figure 5).

**Figure 5:**
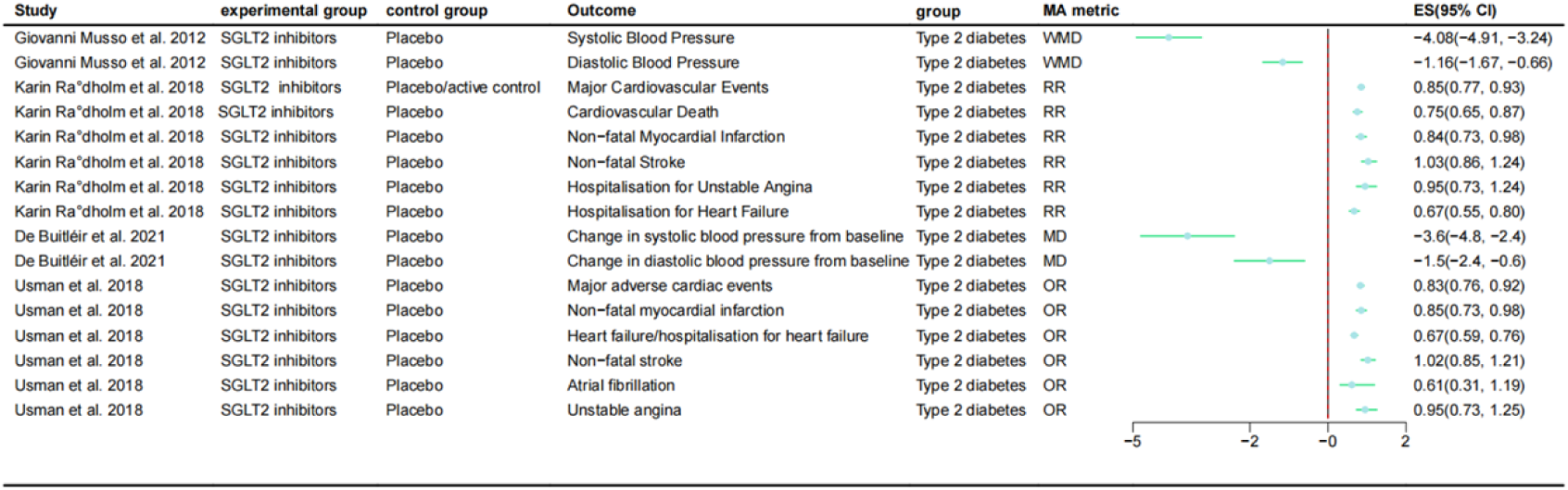
Forest Plot of the Effects of SGLT2 on Cardiovascular Outcomes and Blood Pressure

### 2.5 Hematological Parameters

SGLT2 inhibitors affect certain hematological parameters. The weighted mean difference (WMD) in serum uric acid levels was −41.50 μmol/L (95% CI: −47.22 to −35.79). The mean difference (MD) in hemoglobin levels was 5.60 g/L (95% CI: 3.73 to 7.47); the MD in hematocrit levels was 1.32% (95% CI: 1.21 to 1.44). The standardized mean difference (SMD) in ferritin change was −1.15 (95% CI: −2.87 to 0.57), which was not statistically significant (Figure 6).

**Figure 6:**
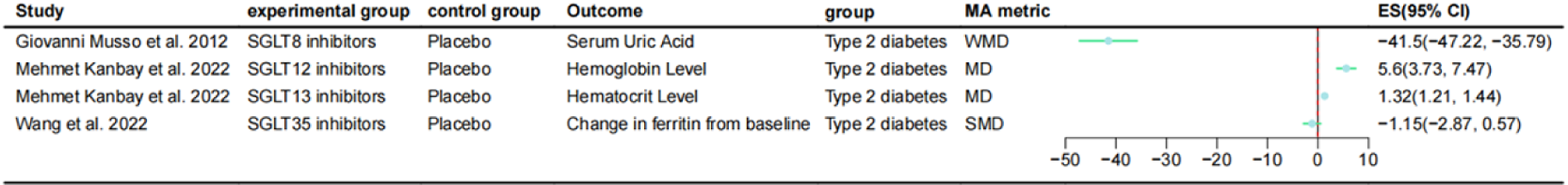
Forest Plot of the Effects of SGLT2 on Hematological Parameters

### 2.6 Safety and Adverse Events

Certain safety indicators of SGLT2 inhibitors warrant attention. The odds ratio (OR) for the occurrence of hypoglycemic events was 1.27 (95% CI: 1.05 to 1.53). The odds ratio (OR) for urinary tract infections was 1.34 (95% CI: 1.05 to 1.71); the odds ratio (OR) for genital tract infections was 3.57 (95% CI: 2.59 to 4.93) (Figure 7).

**Figure 7:**
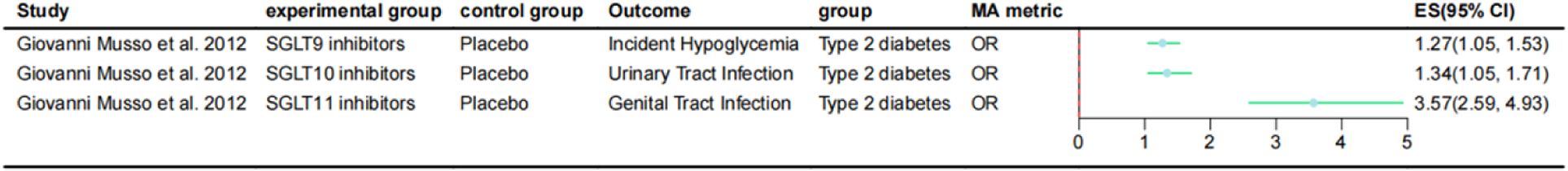
Forest Plot of the Effects of SGLT2 on Safety and Adverse Events

### 2.7 Inflammatory and Adipokine Biomarkers

SGLT2 inhibitors have a regulatory effect on certain inflammatory and adipokine factors. The standardized mean difference (SMD) for changes in C-reactive protein was −0.25 (95% CI: −47 to −0.03); the standardized mean difference (SMD) for changes in tumor necrosis factor-α was −0.05 (95% CI: −0.35 to 0.26), not statistically significant; the standardized mean difference (SMD) for changes in interleukin-6 was −0.57 (95% CI: −1.36 to 0.22), not statistically significant. The standardized mean difference (SMD) for changes in leptin was −0.22 (95% CI: −0.43 to −0.01); the standardized mean difference (SMD) for changes in adiponectin was 0.28 (95% CI: 0.15 to 0.41) (Figure 8).

**Figure 8:**
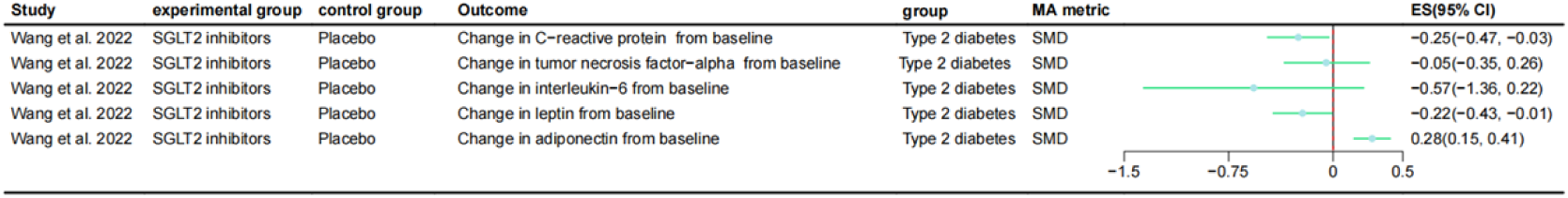
Forest Plot of the Effects of SGLT2 on Inflammatory and Adipokine Biomarkers

### 2.8 Renal Outcomes

SGLT2 inhibitors have shown significant effects in renal protection. The risk ratio (RR) for composite renal outcomes was 0.59 (95% CI: 0.49 to 0.71) (additionally, the RR for composite renal outcomes was 0.64, 95% CI: 0.58 to 0.72; the OR for composite renal outcomes was 0.48, 95% CI: 0.40 to 0.59; the RR for composite renal outcomes was 0.61, 95% CI: 0.45 to 0.82). The RR for progression of albuminuria was 0.72 (95% CI: 0.67 to 0.77); the RR for progression to macroalbuminuria was 0.79 (95% CI: 0.62 to 1.00); the RR for a reduction in estimated glomerular filtration rate (eGFR) ≥40% was 0.62 (95% CI: 0.50 to 0.77) (additionally, the RR for eGFR decline ≥ 40% was 0.58, 95% CI: 0.48 to 0.70); the RR for doubling of serum creatinine was 0.67 (95% CI: 0.56 to 0.81); the RR for dialysis or renal replacement therapy was 0.71 (95% CI: 0.59 to 0.86); the RR for end-stage renal disease was 0.70 (95% CI: 0.56 to 0.87) (additionally, the RR for end-stage kidney disease was 0.65, 95% CI: 0.53 to 0.80); the RR for acute kidney injury was 0.79 (95% CI: 0.71 to 0.89) (additionally, the RR for acute kidney injury was 0.75, 95% CI: 0.66 to 0.85; the OR for acute renal failure or injury was 0.77, 95% CI: 0.66 to 0.91); the RR for sustained eGFR <15 ml/min was 0.66 (95% CI: 0.55 to 0.81); the RR for renal death was 0.53 (95% CI: 0.26 to 1.09); the OR for renal impairment was 1.48 (95% CI: 1.07 to 2.04); the MD for change in eGFR was 0.16 (95% CI: −0.83 to 1.14); the SMD for change in urinary albumin-creatinine ratio (UACR) was −0.21 (95% CI: −0.49 to 0.07)(Figure 9).

**Figure 9:**
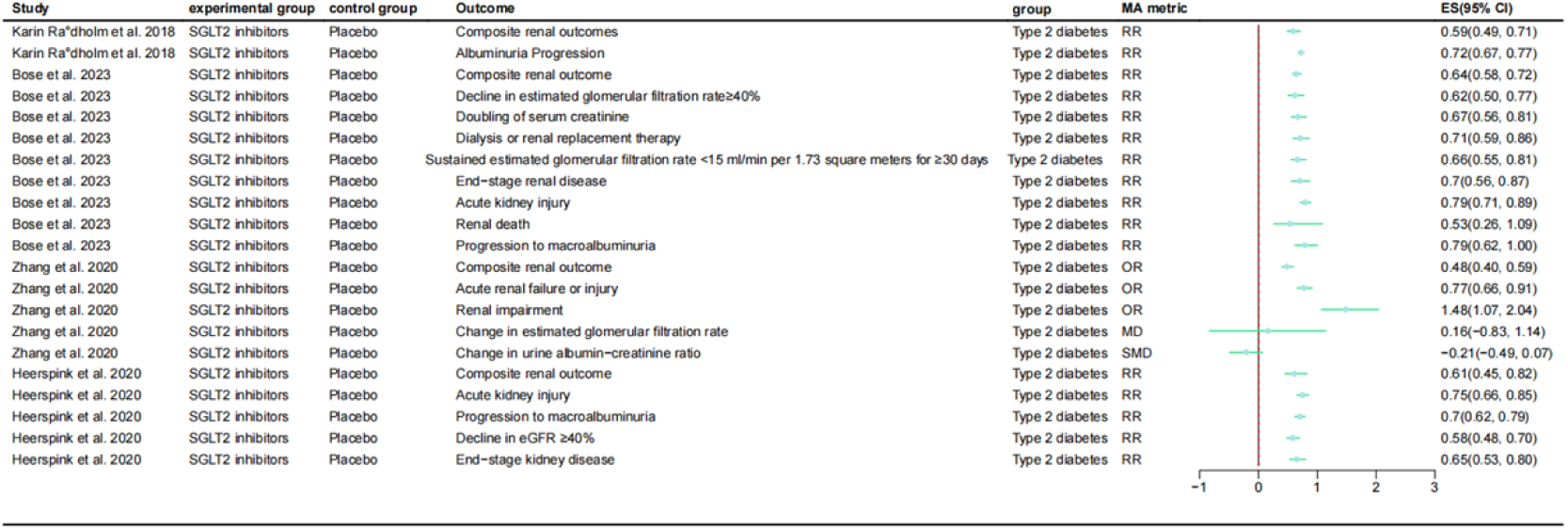
Forest Plot of the Effects of SGLT2 on Renal Outcomes

### 2.9 All-Cause Mortality Rate

SGLT2 inhibitors can significantly reduce all-cause mortality. The risk ratio (RR) for all-cause mortality was 0.79 (95% CI: 0.70 to 0.88); the odds ratio (OR) for all-cause mortality was 0.79 (95% CI: 0.70 to 0.89) (Figure 10).

**Figure 10:**
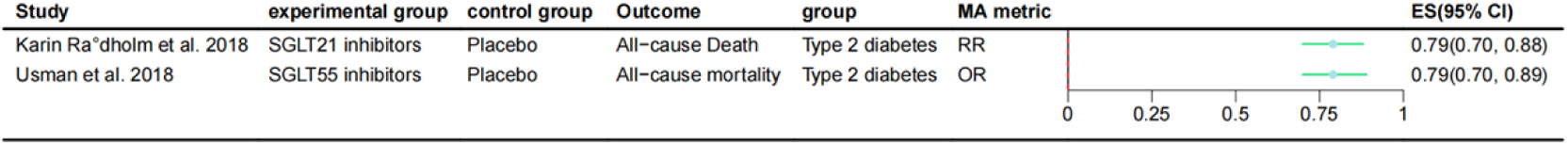
Forest Plot of the Effects of SGLT2 on All-Cause Mortality Rate

### 2.10 Heterogeneity

The heterogeneity of the included studies varied depending on the outcome indicators. Some outcome indicators showed low to moderate heterogeneity, such as glycated hemoglobin (I^2^ = 0.14), fasting blood glucose (I^2^ = 0.18), systolic blood pressure (I^2^ = 0), and several cardiovascular hard endpoints (I^2^ mostly below 0.1). However, some outcome indicators exhibited high heterogeneity, such as body mass index (I ^2^ = 0.9), insulin sensitivity (I ^2^ = 0.848), weight change (I ^2^ = 0.75), and certain inflammatory markers (e.g., change in ferritin I ^2^ = 0.97). The high heterogeneity may stem from differences in intervention protocols, follow-up duration, baseline population characteristics, or measurement methods across studies.

### 2.11 Publication Bias

Egger’s test results showed no significant publication bias (P > 0.05) in most outcome indicators of the meta-analyses. For example, changes in glycated hemoglobin (P > 0.05), changes in fasting blood glucose (P > 0.05), and major cardiovascular events (P > 0.05) all did not indicate the presence of small-study effects. Only a few outcome indicators, such as changes in insulin dosage (P < 0.01) and acute kidney injury (P = 0.029), had Egger’s test P values < 0.1, suggesting possible publication bias. Overall, no significant publication bias was reported in all meta-analyses.

### 2.12 Assessment of Evidence Quality and Risk of Bias

The AMSTAR 2.0 tool was used to assess the methodological quality of the included systematic reviews/meta-analyses. The results showed that most studies performed well in terms of search strategy, bias risk control, and statistical analysis, with the overall quality of evidence rated as moderate to high. The main limitations were that some studies did not have a pre-registered protocol or did not adequately report the list and reasons for excluded studies. The risk of bias assessment indicated that the included original studies were mainly randomized controlled trials, with an overall low risk of bias. Sensitivity analysis results showed that after excluding low-quality studies, the pooled effect size remained robust, indicating a high level of credibility for the evidence of the main outcome measures (Table 2).

**Table 2.**
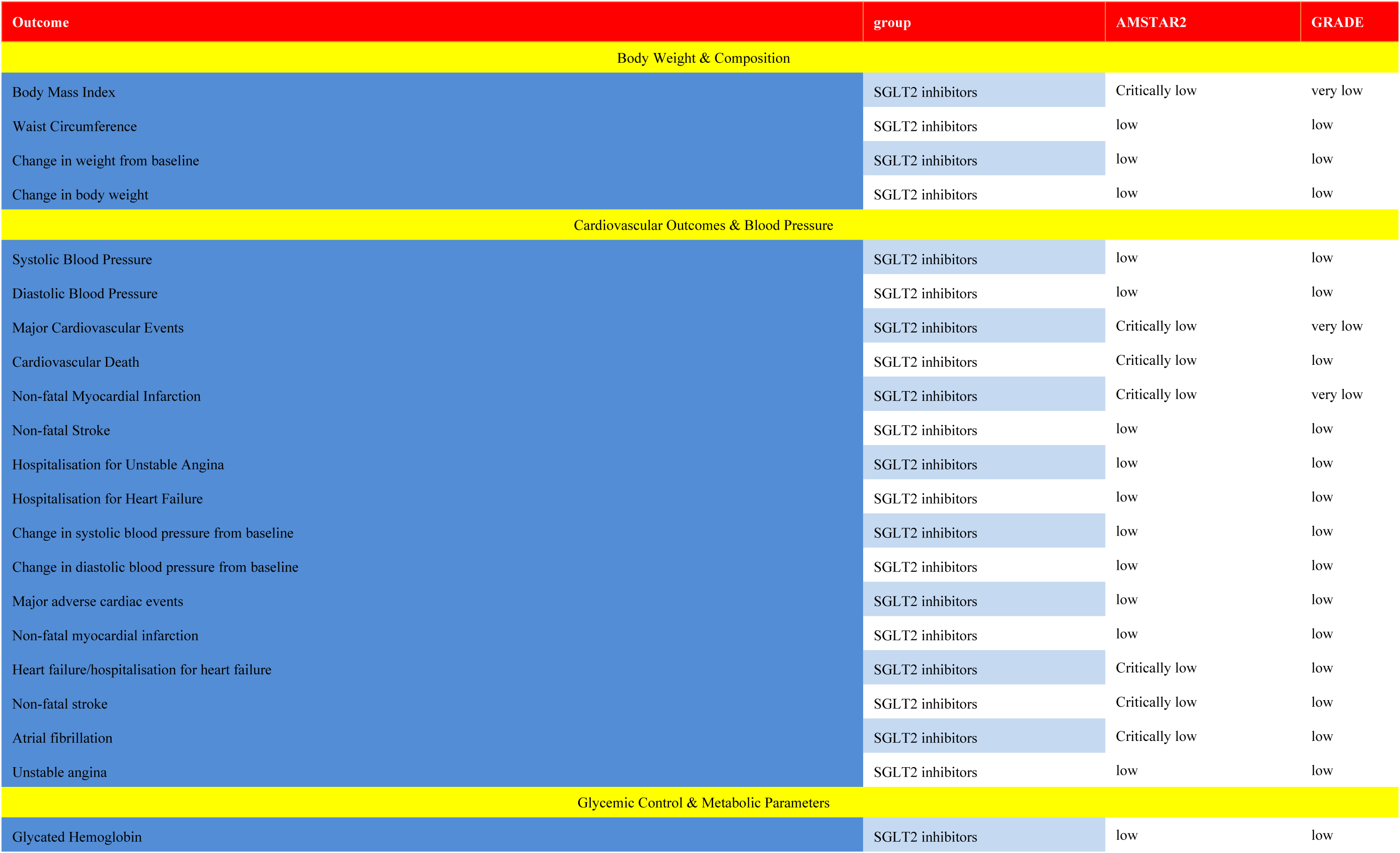
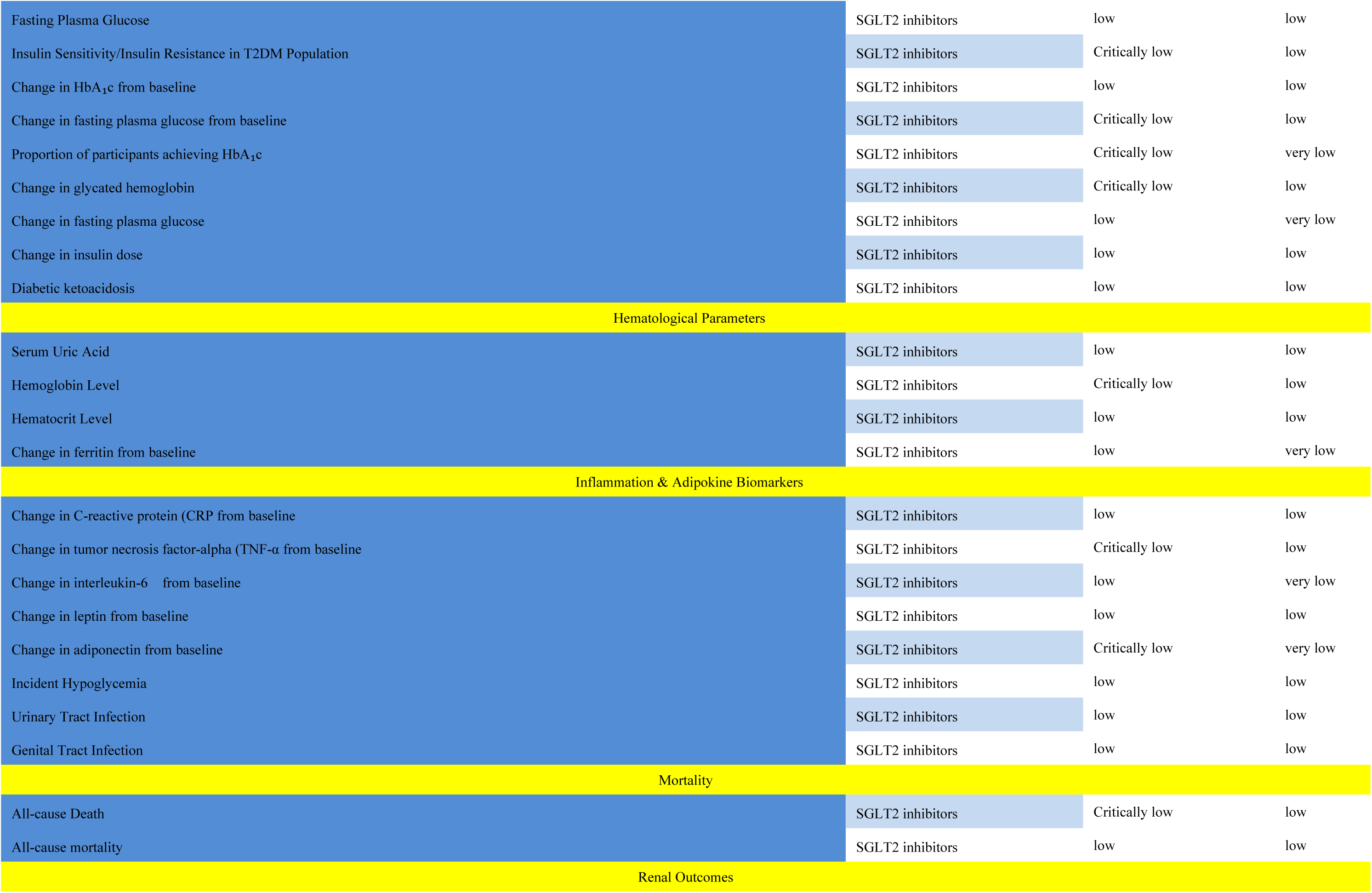
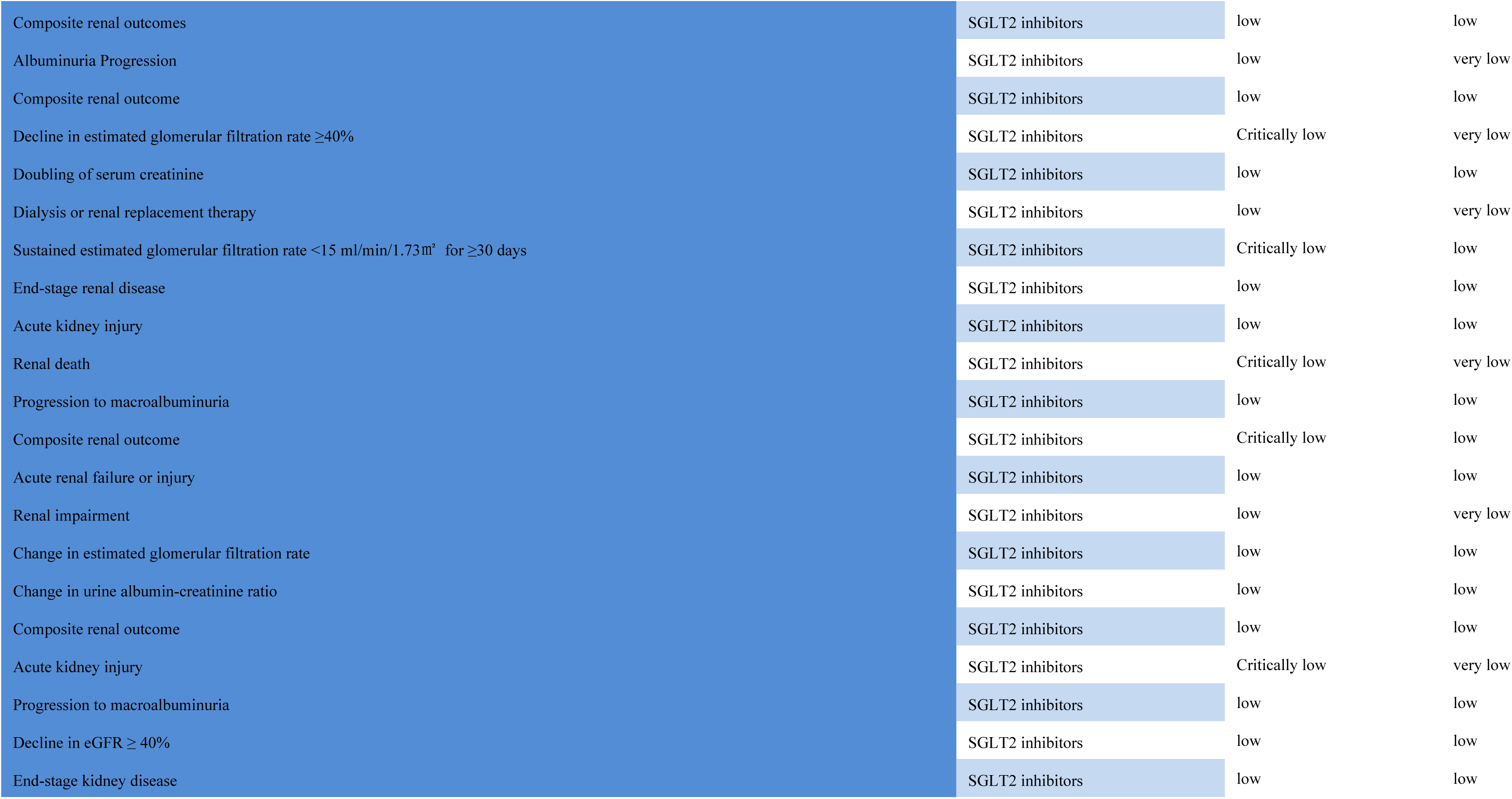
Analysis table of research methodological quality and evidence reliability in SGLT2 based on AMSTAR 2 scores and GRADE grading.

| Outcome | group | AMSTAR2 | GRADE |
| --- | --- | --- | --- |
| Body Weight & Composition |  |  |  |
| Body Mass Index | SGLT2 inhibitors | Critically low | very low |
| Waist Circumference | SGLT2 inhibitors | low | low |
| Change in weight from baseline | SGLT2 inhibitors | low | low |
| Change in body weight | SGLT2 inhibitors | low | low |
| Cardiovascular Outcomes & Blood Pressure |  |  |  |
| Systolic Blood Pressure | SGLT2 inhibitors | low | low |
| Diastolic Blood Pressure | SGLT2 inhibitors | low | low |
| Major Cardiovascular Events | SGLT2 inhibitors | Critically low | very low |
| Cardiovascular Death | SGLT2 inhibitors | Critically low | low |
| Non-fatal Myocardial Infarction | SGLT2 inhibitors | Critically low | very low |
| Non-fatal Stroke | SGLT2 inhibitors | low | low |
| Hospitalisation for Unstable Angina | SGLT2 inhibitors | low | low |
| Hospitalisation for Heart Failure | SGLT2 inhibitors | low | low |
| Change in systolic blood pressure from baseline | SGLT2 inhibitors | low | low |
| Change in diastolic blood pressure from baseline | SGLT2 inhibitors | low | low |
| Major adverse cardiac events | SGLT2 inhibitors | low | low |
| Non-fatal myocardial infarction | SGLT2 inhibitors | low | low |
| Heart failure/hospitalisation for heart failure | SGLT2 inhibitors | Critically low | low |
| Non-fatal stroke | SGLT2 inhibitors | Critically low | low |
| Atrial fibrillation | SGLT2 inhibitors | Critically low | low |
| Unstable angina | SGLT2 inhibitors | low | low |
| Glycemic Control & Metabolic Parameters |  |  |  |
| Glycated Hemoglobin | SGLT2 inhibitors | low | low |
| Fasting Plasma Glucose | SGLT2 inhibitors | low | low |
| Insulin Sensitivity/Insulin Resistance in T2DM Population | SGLT2 inhibitors | Critically low | low |
| Change in HbA<sub>1c</sub> from baseline | SGLT2 inhibitors | low | low |
| Change in fasting plasma glucose from baseline | SGLT2 inhibitors | Critically low | low |
| Proportion of participants achieving HbA<sub>1c</sub> | SGLT2 inhibitors | Critically low | very low |
| Change in glycated hemoglobin | SGLT2 inhibitors | Critically low | low |
| Change in fasting plasma glucose | SGLT2 inhibitors | low | very low |
| Change in insulin dose | SGLT2 inhibitors | low | low |
| Diabetic ketoacidosis | SGLT2 inhibitors | low | low |
| Hematological Parameters |  |  |  |
| Serum Uric Acid | SGLT2 inhibitors | low | low |
| Hemoglobin Level | SGLT2 inhibitors | Critically low | low |
| Hematocrit Level | SGLT2 inhibitors | low | low |
| Change in ferritin from baseline | SGLT2 inhibitors | low | very low |
| Inflammation & Adipokine Biomarkers |  |  |  |
| Change in C-reactive protein (CRP from baseline | SGLT2 inhibitors | low | low |
| Change in tumor necrosis factor-alpha (TNF-α from baseline | SGLT2 inhibitors | Critically low | low |
| Change in interleukin-6 from baseline | SGLT2 inhibitors | low | very low |
| Change in leptin from baseline | SGLT2 inhibitors | low | low |
| Change in adiponectin from baseline | SGLT2 inhibitors | Critically low | very low |
| Incident Hypoglycemia | SGLT2 inhibitors | low | low |
| Urinary Tract Infection | SGLT2 inhibitors | low | low |
| Genital Tract Infection | SGLT2 inhibitors | low | low |
| Mortality |  |  |  |
| All-cause Death | SGLT2 inhibitors | Critically low | low |
| All-cause mortality | SGLT2 inhibitors | low | low |
| Renal Outcomes |  |  |  |
| Composite renal outcomes | SGLT2 inhibitors | low | low |
| Albuminuria Progression | SGLT2 inhibitors | low | very low |
| Composite renal outcome | SGLT2 inhibitors | low | low |
| Decline in estimated glomerular filtration rate $\geq 40\%$ | SGLT2 inhibitors | Critically low | very low |
| Doubling of serum creatinine | SGLT2 inhibitors | low | low |
| Dialysis or renal replacement therapy | SGLT2 inhibitors | low | very low |
| Sustained estimated glomerular filtration rate $< 15$ ml/min/1.73m<sup>2</sup> for $\geq 30$ days | SGLT2 inhibitors | Critically low | low |
| End-stage renal disease | SGLT2 inhibitors | low | low |
| Acute kidney injury | SGLT2 inhibitors | low | low |
| Renal death | SGLT2 inhibitors | Critically low | very low |
| Progression to macroalbuminuria | SGLT2 inhibitors | low | low |
| Composite renal outcome | SGLT2 inhibitors | Critically low | low |
| Acute renal failure or injury | SGLT2 inhibitors | low | low |
| Renal impairment | SGLT2 inhibitors | low | very low |
| Change in estimated glomerular filtration rate | SGLT2 inhibitors | low | low |
| Change in urine albumin-creatinine ratio | SGLT2 inhibitors | low | low |
| Composite renal outcome | SGLT2 inhibitors | low | low |
| Acute kidney injury | SGLT2 inhibitors | Critically low | very low |
| Progression to macroalbuminuria | SGLT2 inhibitors | low | low |
| Decline in eGFR $\geq 40\%$ | SGLT2 inhibitors | low | low |
| End-stage kidney disease | SGLT2 inhibitors | low | low |

## 3 Conclusion

In summary, this umbrella review demonstrates that SGLT2 inhibitors offer multiple benefits in patients with T2DM, including glycemic control, weight reduction, and cardiorenal protection. This provides a scientific basis for their clinical application and the development of personalized treatment strategies. Additionally, the review suggests the need for further exploration of efficacy differences across various eGFR stages and levels of proteinuria, as well as comparisons of efficacy and safety among different types of SGLT2 inhibitors.

## Data Availability

The data underlying this umbrella review are derived from published systematic reviews and meta-analyses, which can be retrieved from public databases including Embase, Medline, the Cochrane Database of Systematic Reviews, and Web of Science. Supporting analytical data can be obtained from the corresponding author, Rui Ma, via email at upon reasonable request.

